# Lessons learned from real-time nowcasting: The 2024 dengue outbreak in Puerto Rico

**DOI:** 10.64898/2026.07.20.26358497

**Authors:** Quan M Tran, Ariana Detmar, Carol Y Liu, Zachary J Madewell, Dania M Rodriguez, Jomil Torres Aponte, Melissa Marzán-Rodriguez, Gabriela Paz-Bailey, Laura E Adams, Karen Holcomb, Michael A Johansson, Maile B Thayer

## Abstract

Real-time nowcasting enhances situational awareness by mitigating reporting delays that obscure transmission dynamics. We applied Nowcasting by Bayesian Smoothing (NobBS) to the 2024 dengue outbreak in Puerto Rico (PR), using case surveillance data from the PR Department of Health. The method accurately captured the epidemic trajectory and consistently outperformed a baseline model, although reporting anomalies occasionally reduced performance. We also conducted analyses by dengue virus serotype and health region, as well as previous years. For analyses with few dengue cases, a model in which parameters are jointly estimated across groups generally achieved better performance than the independent one. Historical analyses revealed that years with higher variability in reporting delays generally exhibited higher uncertainty. The findings here underscore key lessons for real-time dengue nowcasting: alternative models may be needed in complex circumstances, but with stable reporting patterns and continuous evaluation, nowcasts can be a reliable and valuable public health tool.

## Introduction

Timely and accurate situational awareness is essential for effective outbreak response (Jajosky and Groseclose 2004). Case surveillance is central in monitoring infectious disease trends and informing public health decision making. However, reporting delays occur frequently due to lags between symptom onset and care-seeking, time needed for clinical or laboratory confirmation, and the confirmation-to-reporting interval (Swaan et al. 2019). As a result, reported cases often actually occurred days or weeks earlier, and do not represent real-time transmission levels. When cases are shown in epidemiologic curves by symptom onset dates, recent counts may appear artificially low, creating the false perception that transmission is declining. These delays complicate interpretation of the current outbreak status and may hinder critical response activities (Swaan et al. 2019; Salmon et al. 2015).

To address these limitations, nowcasting methods have been developed to estimate the true number of cases that have occurred but have not yet been reported. Nowcasting enables more accurate real-time assessment of disease activity by accounting for reporting delays, supporting more timely and effective public health response. A variety of nowcasting methods have been applied in outbreak contexts, ranging from simple delay adjustment models to fully Bayesian approaches (Noufaily et al. 2015; McGough et al. 2020; Charniga et al. 2024).

We applied a nowcasting for the three most recent weeks to dengue in Puerto Rico, similar to what has been implemented in Brazil (Codeco et al. 2018). Dengue is a mosquito-borne viral disease caused by dengue virus (DENV) that has four distinct serotypes (DENV-1 to DENV-4) and can cocirculate in tropical and subtropical regions (Paz-Bailey et al. 2024). It is endemic in Puerto Rico, with periodic outbreaks, most notably in 2010 and 2013 (Sharp et al. 2019). Shortly after nowcasting was implemented in the surveillance system in early 2024, the system detected a rise in nowcasted cases that surpassed the epidemic threshold in late January, ultimately contributing to a declaration of a public health emergency by the Puerto Rico Department of Health (PRDH) (Ware-Gilmore et al. 2025; Puerto Rico Department of Health 2024).

Here, we describe the application and insights gained from the implementation of the nowcasting system during the 2024 dengue outbreak in Puerto Rico. We also apply the same framework to create nowcasts by health region and DENV serotype, and retrospectively to prior outbreaks and interepidemic periods to assess their performance across time.

## Materials and Methods

### Data

To generate nowcast estimates, we used historical dengue case data from the Puerto Rico Passive Arboviral Disease Surveillance System (PADSS), which includes all probable and confirmed DENV cases from January 1, 1986, to December 31, 2024. Probable cases are defined by a positive immunoglobulin M (IgM) antibody test for DENV, while confirmed cases have a positive result for DENV by reverse transcription polymerase chain reaction (RT-PCR). For this analysis, we aggregated case data by epidemiological week (EW) and year. For each case, we assessed both the date of the patient’s symptom onset and the date of reporting, corresponding to when the case was entered into PADSS. Probable and confirmed cases were modeled jointly for all analyses, with a single delay distribution estimated across both case types.

Previous work has established an epidemic alert threshold by fitting historical case data to an intercept-only negative binomial regression model (Thayer et al. 2024). This threshold has been used to monitor reported cases and signal when case counts surpass levels indicative of an outbreak. In this study, we extend its application by monitoring nowcasted case estimates against the threshold, enabling earlier detection and improving situational awareness.

### Model

We used Nowcasting by Bayesian Smoothing (NobBS), a flexible Bayesian framework that models the delay distribution between symptom onset and report date using historical case line data (McGough et al. 2020). NobBS model includes four key parameters: The time-varying epidemiological signal (*α_t_*) of dengue incidence at time *t* and its variance (*φ*), the reporting delay probabilities (*β_d_*) at week *d*, and the dispersion parameter (*r*) of case counts. NobBS does not require disease-specific parameterization, making it broadly applicable across different pathogens. It was previously applied to real-time surveillance of diseases such as COVID-19, influenza-like illness, and mpox (McGough et al. 2020; Lopez et al. 2024; Charniga et al. 2024). Here, we used a modified version of NobBS, reimplemented in Stan to improve computational performance (Stan Development Team 2022).

We generated weekly real-time nowcasts throughout the dengue outbreak in 2024, estimating 50% and 95% predictive intervals (PIs) for two weeks prior, one week prior, and the current week, denoted as horizons -2, -1, and 0 respectively. Nowcasts were created at the island-wide level, as well as by health region (n=8) and DENV serotype (n=4) to more closely track spatial patterns and DENV serotypes, supporting a more targeted outbreak response. The eight health regions were defined by PRDH to address specific geographic needs and centralize healthcare services (Stimpson et al. 2024).

For the nowcasting by health region and DENV serotype, we fitted two types of models: one where all parameters were unique to each subset group and one with some shared parameters among the group. Unique parameters capture differences in transmission (*α_t_*) and reporting (*β_d_*) across groups, while shared parameters capture the uncertainty (*r* and *φ*). This pooling leveraged information from high-case-count groups to stabilize estimates in low-case-count groups, improving statistical power and precision of inferences.

We retrospectively applied the nowcasting framework to historical data, by performing weekly nowcasts using only the data reported up to each respective week, to evaluate applicability of our nowcasting framework across diverse epidemic conditions. Specifically, we selected two large historical outbreak years in Puerto Rico (2010 and 2013), as well as the inter-epidemic year (2009), to assess performance under both high- and low-incidence scenarios.

A detailed description of the models is in the Appendix, along with our modifications applied for alternative models. The R code for this analysis is available on GitHub (Codes).

### Model assessment

To benchmark whether a more complex nowcasting method like NobBS provides meaningful improvements, we used a simple baseline model where the predicted mean case count equals to the most recent observed value in the week before the nowcasting window, with the variability derived from historical week-to-week differences in case counts. This baseline model is previously described in Cramer et al. (Cramer et al. 2022) and further outlined in the Appendix.

Model performance was evaluated using weighted interval scores (WIS), (Bracher et al. 2021), a proper scoring rule that balances accuracy and uncertainty. Lower WIS values indicate better performance. To better understand model performance, we decomposed the WIS into four components: dispersion and absolute error, which reflect the prediction uncertainty and accuracy respectively, and underprediction and overprediction, which measure the penalties for predictions falling above and below the observed value (see Appendix for details).

We additionally evaluated the coverage of the models’ 50% and 95% PIs to assess model calibration, with well-calibrated intervals expected to contain eventually reported cases approximately 50% and 95% of the time, respectively. To capture rare but consequential failures, we defined a failed nowcast as one with a log score below -9.2 (probability ≤ 1 in 10,000) for at least one of the three most recent weeks. This threshold identifies nowcasts with exceptionally low predictive probabilities (Lopez et al. 2024) and is slightly more stringent compared to the -10 used in the CDC FluSight Challenge (Reich et al. 2019).

## Results

### Nowcasting in 2024

#### Dengue outbreak detection in Puerto Rico

We implemented nowcasting for Puerto Rico in January 2024, prior to the outbreak start (Fig 1). Although case counts are typically low in January 6^th^, island-wide nowcasts signaled that dengue case numbers could be nearing the epidemic threshold. On January 27^th^, the 25^th^ percentile of the nowcast exceeded the epidemic threshold while reported cases remained below it, indicating at least a 75% probability of elevated transmission. On February 3^rd^, reported cases first exceeded the epidemic threshold, about four weeks after the nowcast cases did on January 6^th^, providing valuable early warning time before the outbreak. By February 17^th^, the lower bound of the 95% PI exceeded the threshold, indicating a greater than 95% probability that cases were above the epidemic threshold. From that point onward, nowcasts generally remained above the threshold, contributing to the formal declaration of a public health emergency in Puerto Rico in March.

**Figure 1.**
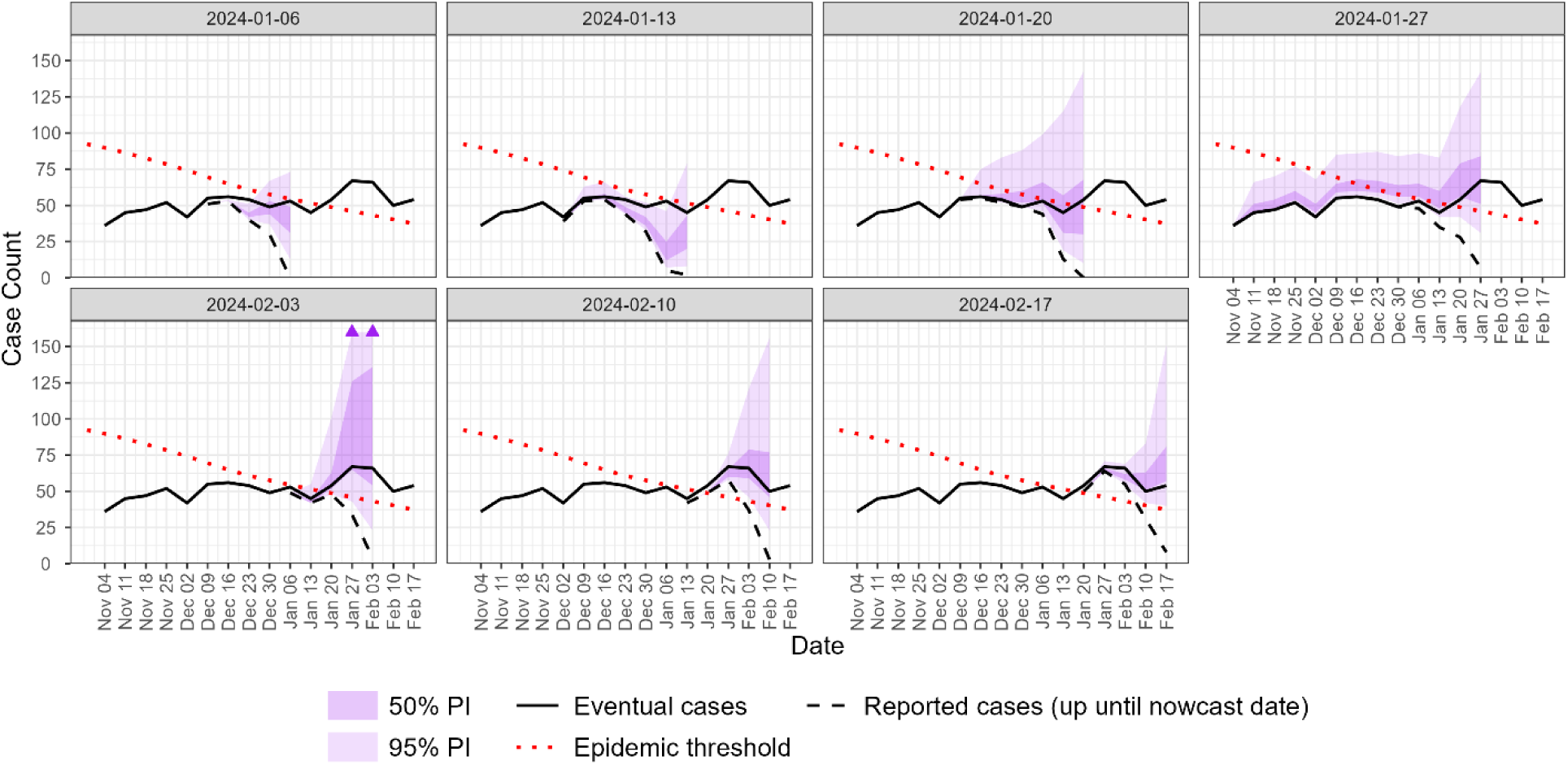
Real-time island-wide nowcasts across the first six weeks of 2024 in Puerto Rico (January 6 to February 10). Each panel corresponds to a distinct nowcast date and shows reported cases as of that date (dashed black line), eventual case counts (solid black line), and the predefined epidemic threshold (dotted red line). Shaded regions represent 50% (light purple) and 95% (dark purple) predictive intervals from the NobBS model. Triangles indicate 95% prediction interval upper bounds exceeding 160 cases. Case counts are plotted on a logarithmic scale. PI = nowcast prediction interval.

#### Island-wide nowcasts in 2024

Weekly island-wide nowcasts generally predicted the final case numbers that were eventually reported in 2024, capturing both the overall increase in incidence and the timing of the peak several weeks ahead of reported case numbers (Fig 2). For most of the year, nowcast estimates remained above the epidemic threshold, as expected during an ongoing epidemic. Under the definition of failed nowcast criteria (log-score < -9.2 in at least one of the three horizons), EWs 39, 43, 45, and 47 were flagged as failed nowcasts. We noticed a delayed batch reporting event occurred in EW 47, during which 119 of total 304 reported cases for the week had delays, exceeding four weeks and spanning up to three months prior. This worsened the nowcasts performance before and at EW 47. If we truncate the case line to reflect data available at that time (i.e., ignore the batch reporting and use only cases reported as of EW 47) and reevaluate nowcast performance, EWs 39, 43, and 45 are no longer classified as failures. For EW 47, it should be interpreted with caution due to the mentioned reporting anomaly.

**Figure 2.**
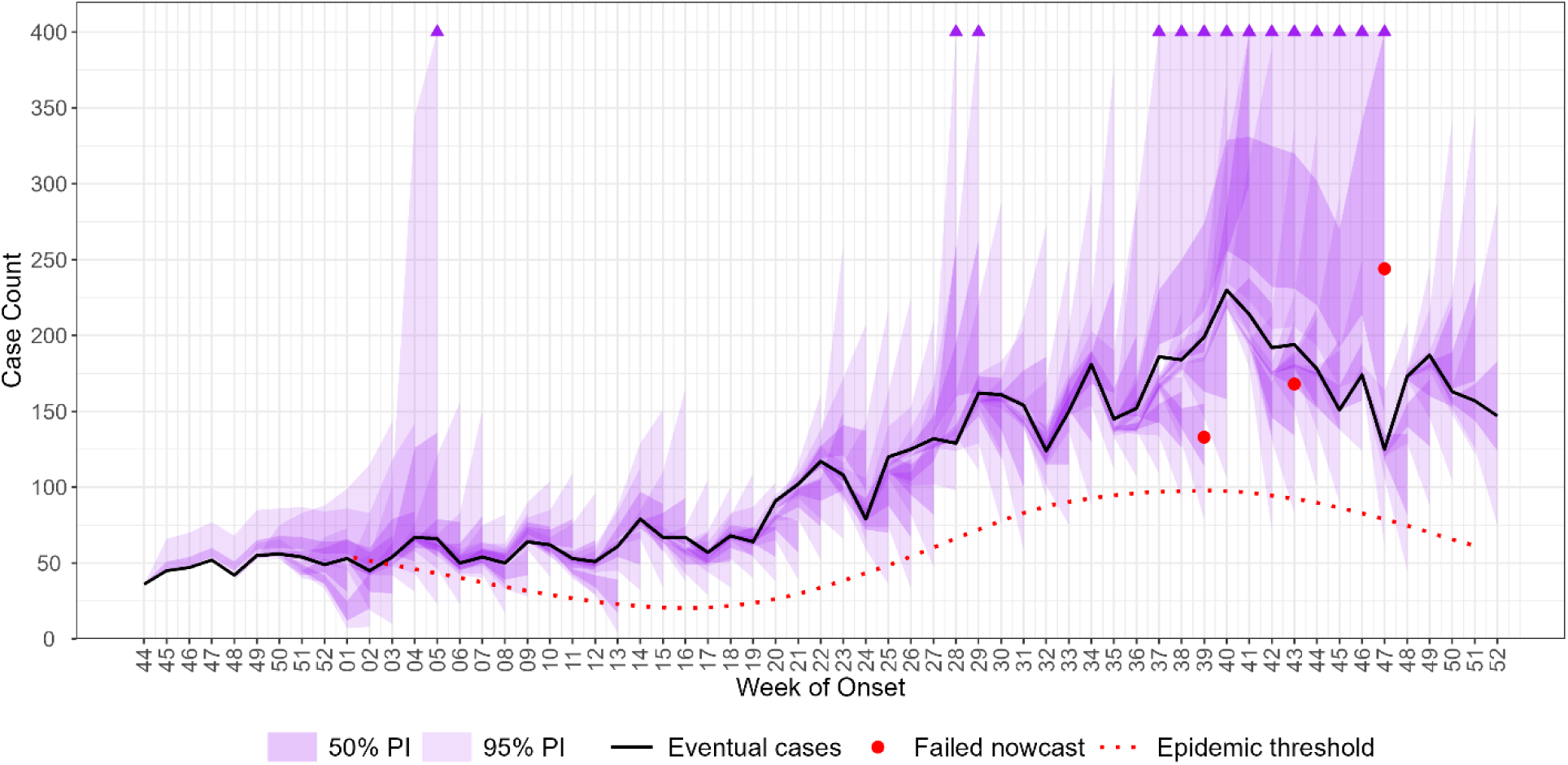
Overlaying real-time island-wide nowcasts across 2024 in Puerto Rico. The graph shows the eventual case counts (solid black line) and the predefined epidemic threshold (dotted red line). Shaded regions represent 50% (light purple) and 95% (dark purple) predictive intervals from the NobBS model. Red dots mark failed nowcasts in EW 38, 42, 44, 46, defined by log-score. Triangles indicate 95% prediction interval upper bounds exceeding 400 cases. Case counts are plotted on a logarithmic scale. PI = nowcast prediction interval.

Across all horizons and EWs in 2024, the median coverage was 0.37 (0.30-0.45) for the 50% PI and 0.88 (0.82-0.93) for the 95% PI, which are slightly below the 50% and 95% targets (Appendix Table). This indicates that the eventual reported dengue counts fell within prediction intervals less frequently than expected and suggests that PIs were somewhat narrow. Coverage improved to 0.40 (0.33-0.48) and 0.89 (0.83-0.94) respectively when excluding the EW 47 batch. The median coverage for horizon 0 across EWs, which is usually the most valuable in real-time surveillance, had excellent coverage (median 0.48 (0.35-0.62) for the 50% PI and 1.00 (1.00-1.00) for the 95% PI).

In the 2024 island-wide nowcasts, the NobBS model yielded a WIS of 76.7, with dispersion (48.6), followed by overprediction (18.6), underprediction (6.2), and absolute error (3.3). Overprediction exceeded underprediction, indicating a tendency to overestimate cases. In contrast, the baseline model had a higher WIS (134.1), higher dispersion (104.4), and greater absolute error (6.2). Therefore, NobBS outperformed the baseline across all components (except overprediction), substantially reducing error and improving precision.

#### Nowcasts by region and DENV serotype in 2024

Nowcasts by health region and DENV serotype showed strong overall performance in 2024, with both models closely tracking final reported case numbers (Fig. 3 and 4). When case counts were low in certain regions, the shared parameter model produced predictions with lower uncertainty compared to the independent parameter model. This reduced uncertainty occurs because the shared parameter approach pools information across regions, stabilizing estimates and reducing uncertainty, particularly in regions with low case count.

**Figure 3.**
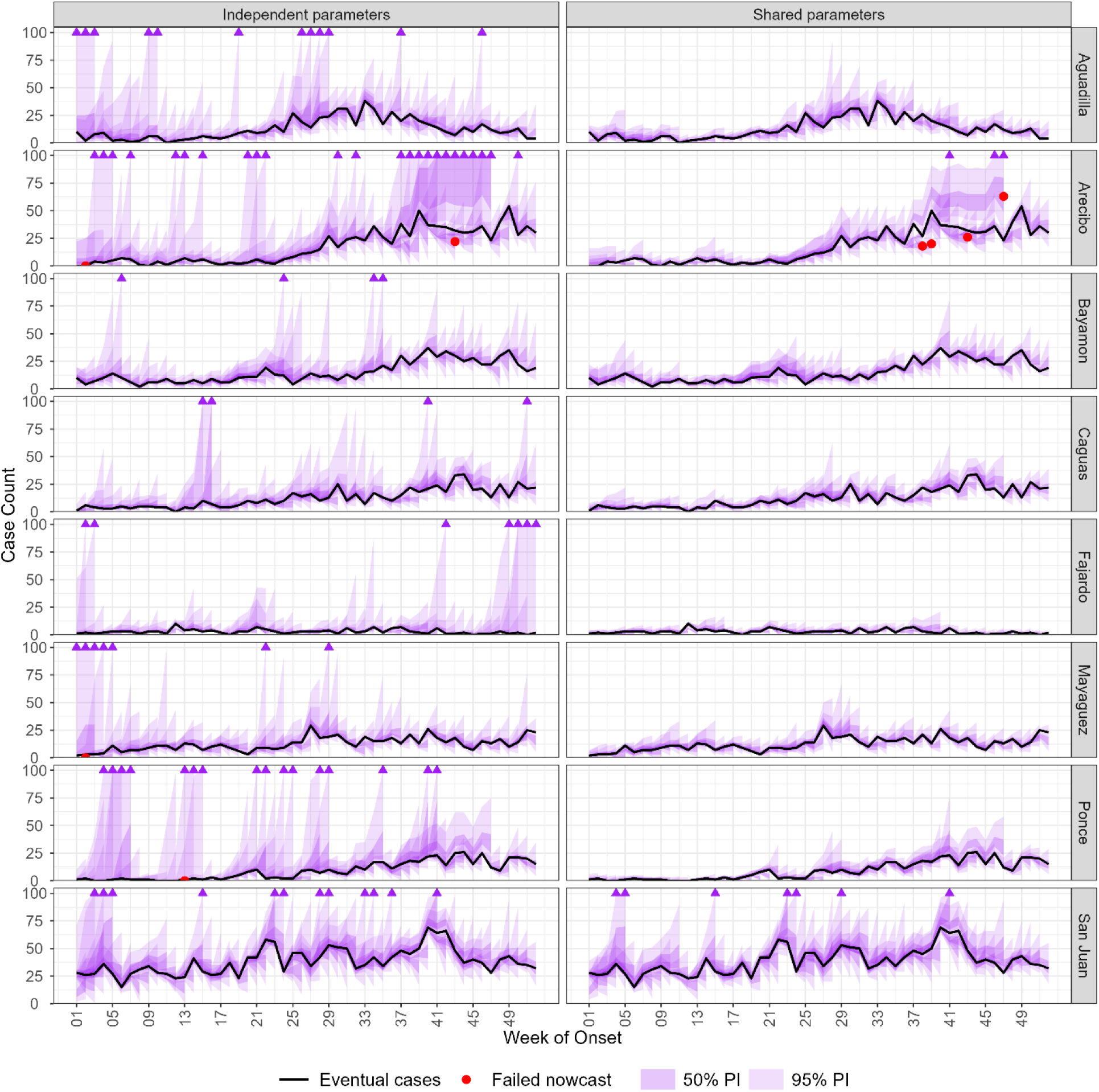
Overlaying real-time nowcasts for each health region across 2024 in Puerto Rico. The graph shows the eventual case counts (solid black line) and the predefined epidemic threshold (dotted red line). Shaded regions represent 50% (light purple) and 95% (dark purple) predictive intervals from models where all parameters are independent (left) or some parameters are shared (right). Red dots mark failed nowcasts defined by log-score. Triangles indicate 95% prediction interval upper bounds exceeding 100 cases. Case counts are plotted on a logarithmic scale. PI = nowcast prediction interval.

**Figure 4.**
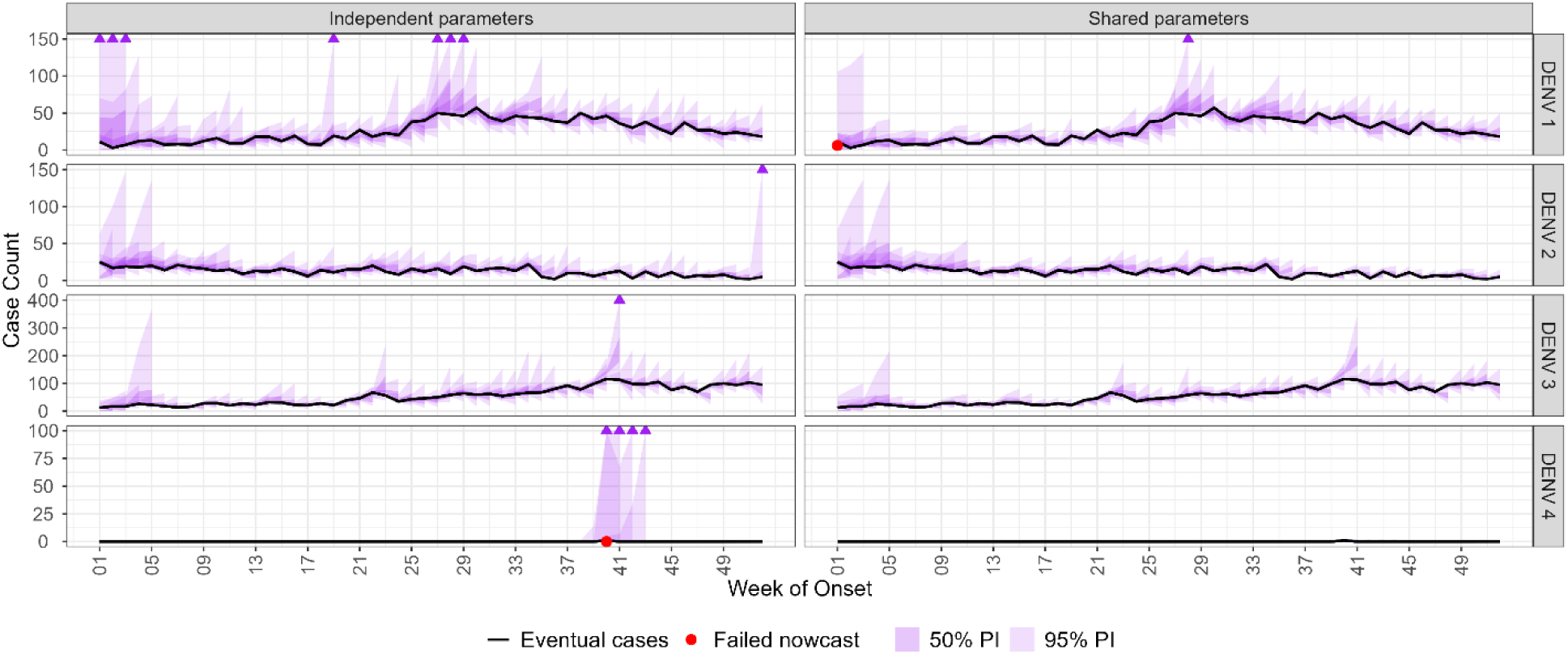
Overlaying real-time nowcasts for each serotype across 2024 in Puerto Rico. The graph shows the eventual case counts (solid black line) and the predefined epidemic threshold (dotted red line). Shaded regions represent 50% (light purple) and 95% (dark purple) predictive intervals from models where all parameters are independent (left) or some parameters are shared (right). Red dots mark failed nowcasts defined by log-score. Triangles indicate 95% prediction interval upper bounds exceeding 150 cases for DENV 1, 150 cases for DENV 2, 400 cases for DENV 3, and 100 cases for DENV 4. Case counts are plotted on a logarithmic scale. PI = nowcast prediction interval

Several failed nowcasts occurred in multiple regions between EWs 38 and 47 for both models (Fig. 4). The major batch reporting event in EW 47 was traced to the Arecibo health region and was not observed elsewhere. Additional failed nowcasts occurred in the independent parameter model during periods of very low case counts (e.g., EW 2 in Arecibo and Mayagüez, EW 13 in Ponce). Conversely, some EWs showed failed nowcasts in the shared parameter model but not in the independent parameter model (e.g., EWs 38, 39, and 47 in Arecibo), likely reflecting the trade-off between flexibility and pooling. The independent parameter model better adapts to small populations by producing wider prediction intervals that capture reported cases. Tighter intervals produced by the shared parameter model can be overconfident when local patterns deviate from the pooled trend.

Serotype-specific nowcasts showed similar overall performance to regional models, accurately tracking reported cases (Fig. 3). Like regional analyses, the independent parameter model produced higher uncertainty, particularly when case counts were low. Notably, only a single DENV-4 case was reported in EW 40 for all of 2024, in a patient with recent travel outside of Puerto Rico. Given such an extremely low incidence, the independent parameter model generated very wide prediction intervals, which contributed to its failure under our criteria, whereas the shared parameter model did not exhibit this issue. Unlike the regional analysis, the EW 47 batch reporting event had no impact on serotype-specific nowcasts, because the backlog lacked serotype information and was excluded from the analyses.

Coverage of the 50% and 95% PIs in 2024 was generally close to their targets for both health region- and serotype-specific nowcasts (Appendix Fig. 1). Some low-incidence regions, such as Caguas, Fajardo, and Ponce, as well as DENV-2, showed high 50% PI coverage, indicating wider-than-expected intervals. Differences in coverage between the independent and shared parameter models were minimal.

We compared WIS across the independent parameter, shared parameter, and baseline models (Fig 5). In general, the shared parameter model achieved the lowest WIS, indicating the best performance for both health region and serotype-specific data. In contrast, the independent parameter model performed worse than the baseline in some cases, with the WIS exceeding baseline values at certain horizons (for example, horizon −1 and 0 for health region data, and horizon 0 for serotype specific data).

**Figure 5.**
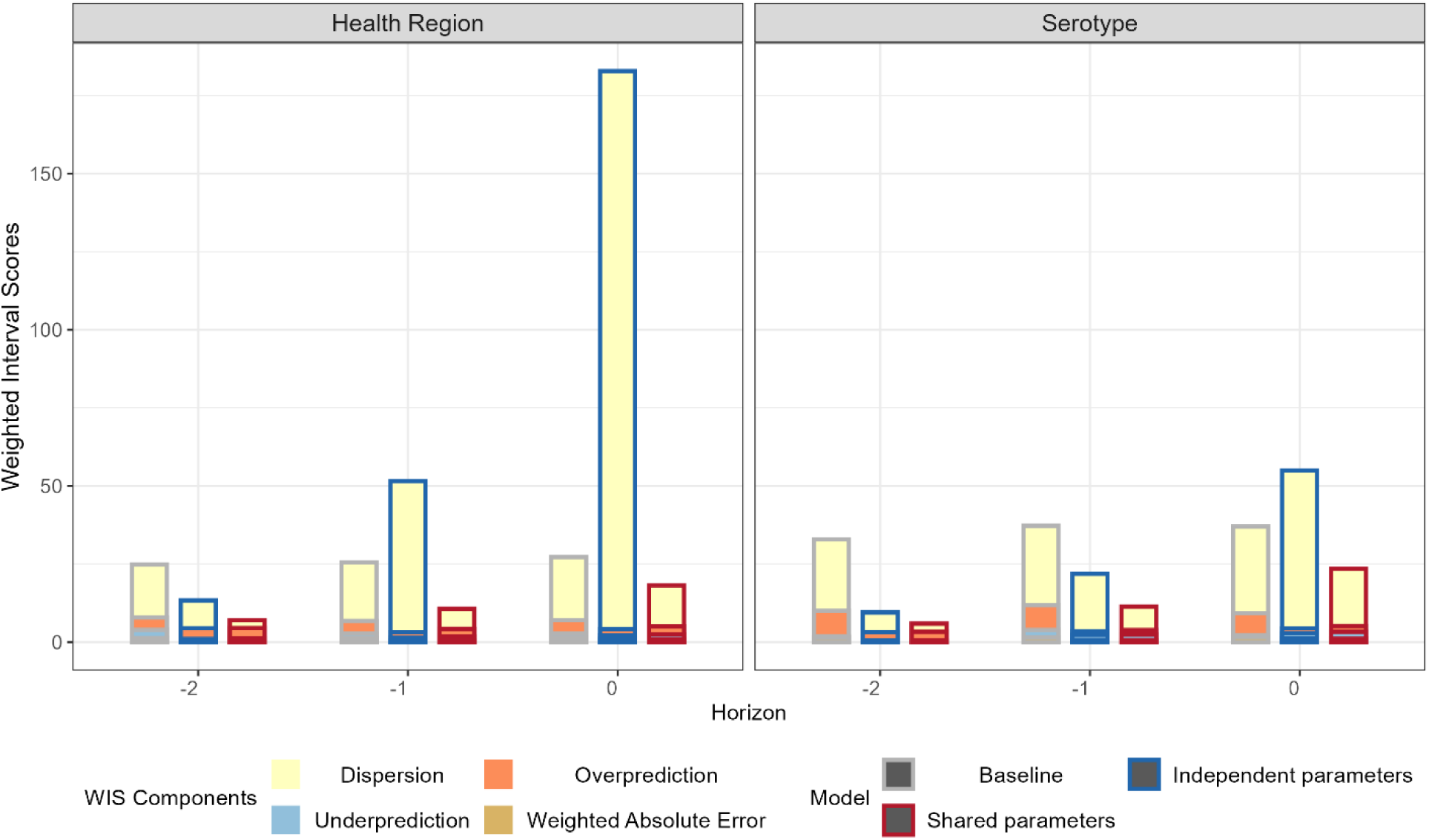
Decomposition of Weighted Interval Score (WIS) for dengue nowcasts in Puerto Rico by health region (left) and serotype (right) across three temporal horizons (−2, −1, 0 weeks). Each bar displays the additive components of WIS filled with different colors: dispersion (yellow), overprediction (orange), underprediction (blue). Independent parameter models (blue), shared parameter models (red), and baseline model (gray) are compared across health regions and serotypes. Lower bar height indicates better model performance by WIS.

WIS decomposition revealed that the independent parameter model had the largest dispersion, reflecting wider PIs (Fig 5). Despite this, the sum of the remaining components was lower for both independent and shared parameter models relative to the baseline, confirming the overall advantage of these models. Notably, the shared parameter model exhibited slightly larger underprediction and overprediction components than the independent parameter model, suggesting reduced ability to capture local variation due to pooling.

To assess the impact of the number of cases on model performance, we examined the relationship between WIS and mean cases across subpopulation (Appendix Fig. 2). As expected, WIS increased with case counts for the shared parameter and baseline models. In contrast, the independent parameter model showed no clear trend, indicating less stable performance likely driven by higher uncertainty. Decomposition further showed that all WIS components except dispersion increased with case counts (Appendix Fig. 2).

### Nowcasting performance in previous years

Nowcasts for 2009, 2010, and 2013 generally tracked the overall case trajectories well, with 50% and 95% PI coverages aligned well with their targets, and only a few failures across these years (Fig 6, Appendix Fig. 2-5, and Appendix Table). In comparison, the baseline model’s PIs tended to exceed the target coverage levels, implying overly broad intervals. However, the 2013 nowcasts displayed unusually high uncertainty. A sensitivity analysis in this period revealed highly variable reporting delays, with average delays ranging from two to six weeks. This demonstrates that model accuracy worsens during periods of erratic change in reporting delays (Appendix Sensitivity Analysis, Appendix Figure 3 and 4).

**Figure 6.**
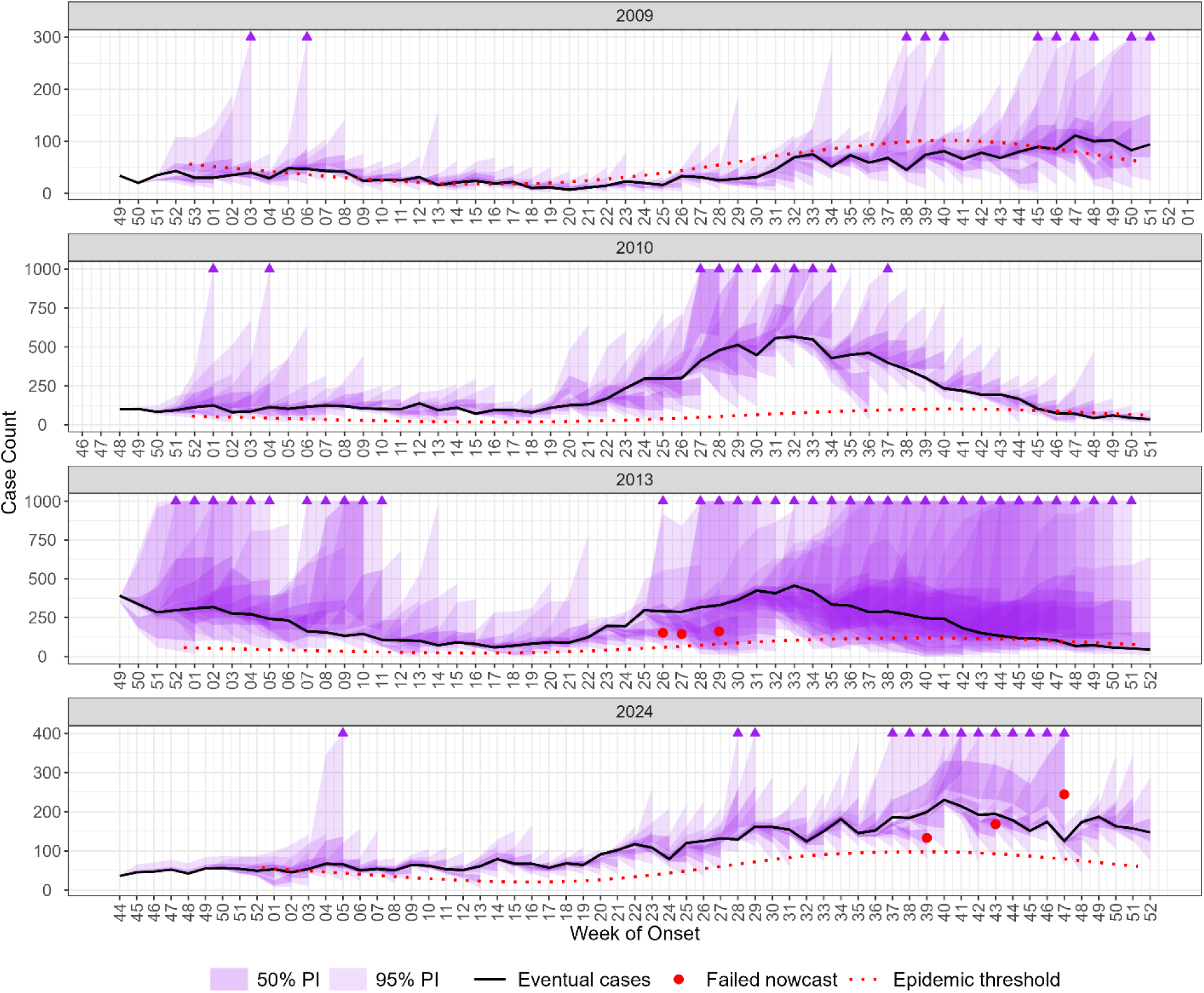
Overlaying real-time nowcasts island-wide for 2009, 2010, 2013, and 2024 in Puerto Rico. The graph shows the eventual case counts (solid black line) and the predefined epidemic threshold (dotted red line). Shaded regions represent 50% (light purple) and 95% (dark purple) predictive intervals from the NobBS model. Red dots mark failed nowcasts defined by log-score. Triangles indicate 95% prediction interval upper bounds exceeding 300 cases for 2009, 1000 cases for 2010, 1000 cases for 2013, and 400 cases for 2024. Case counts are plotted on a logarithmic scale. PI = nowcast prediction interval

## Discussion

We established a robust real-time nowcasting pipeline for dengue case trends in Puerto Rico, validated across multiple years, regions, and DENV serotypes. The real-time 2024 nowcasts demonstrated good performance, effectively capturing the epidemic trajectory and providing timely situational awareness to PRDH. Notably, incorporating shared parameter models improved predictive accuracy and uncertainty in the models by regions and serotypes, as pooling information for low case counts resulted in more stable and reliable estimates.

Incorporating nowcasting into the surveillance system provided valuable early warning before and situational awareness during the 2024 dengue outbreak. In January 2024, our nowcast estimates exceeded the epidemic alert threshold four weeks before observed cases reached the threshold. This finding provided actionable evidence of an unusual increase in cases during the traditional low season, which heightened awareness and supported the declaration of a dengue epidemic in Puerto Rico. This declaration increased attention and resources for case surveillance, mosquito surveillance, raising public awareness, and vector control activities (Ware-Gilmore et al. 2025).

Our retrospective analyses revealed notable differences in nowcast performance across years. The model performed well during both the low (2009) and high (2010) incidence years, but not for the 2013 dengue outbreak, with greater uncertainty largely due to substantial variability in reporting delays during that period. Although we used dynamic parameters that can adjust to changing delays (Lopez et al. 2024), they respond only after those changes appear in the data and therefore lag behind when the changes actually occur. The results show that while the model can handle reporting fluctuations, its performance is limited by data quality and consistency, underscoring the need for stable reporting systems with minimal variation in reporting delays.

From our experience implementing real-time dengue nowcasting in Puerto Rico, several key lessons emerged. First, as mentioned above, having a stable reporting system is critical. Any delays or irregularities, such as those observed during the 2013 outbreak or batch reporting in 2024, can substantially reduce nowcast accuracy. To address this, we routinely monitored reporting delays, compared weekly nowcasts against observed trends, and conducted retrospective evaluations to support model calibration. Routine detection of reporting anomalies should be a common practice for a strong nowcast framework. Second, given the dynamic and complex nature of real-time data, adaptive modeling strategies are essential. Under changing reporting conditions, these strategies may include real-time adjustment of delay distributions, recalibration of nowcasting window, or temporary model switching. We also introduced models that pooled information to improve performance in areas with low case counts. Further enhancement could be made by exploring ensemble methods to reduce single-model bias (Reich et al. 2019; Cramer et al. 2022). Third, predictive alerts must be actionable, requiring strong engagement with local health partners who possess the contextual insight and operational capacity to translate predictions into timely interventions. During 2024, we integrated nowcasts into weekly situational reports, contributing to informed public health decision making. In addition, whenever data anomalies were identified, we initiated timely communication with internal and local health partners to clarify the source of the issue.

After the outbreak was declared, the nowcast continued to support public health decision-making by continuously monitoring DENV transmission levels and providing near real-time estimates of case activity. This helped policy makers to assess whether transmission was increasing, decreasing, or remaining stable, which was valuable for resource allocation, healthcare capacity planning, and implementation of control measures. In 2024, nowcasting helped track the sustained increase in dengue cases throughout the year, with estimated incidence consistently remaining above the epidemic threshold, supporting the continuation of outbreak status in Puerto Rico.

While this study focused primarily on case nowcasting, the framework could be used for other epidemiological metrics, including the time varying reproduction number (Rt) and short-term forecasting. By inputting nowcasted incidence estimates along with their uncertainty into Rt calculation, we can better monitor epidemic trends in real time. Moreover, combining nowcasts of cases with Rt estimates also supports short-term forecasting, which has been implemented in seasonal influenza and mpox (Reich et al. 2019; Charniga et al. 2024). Looking ahead, our next phase aims to integrate Rt, specifically estimating delay distribution and Rt in a single framework, as introduced by (Lison et al. 2024).

Our framework could be applied for application to other arboviruses and geographic settings, provided certain conditions are met. It performs best in contexts with continuous and reasonably reliable data flow, which allows the model to generate stable nowcasts. Because computational demands are minimal, the framework can be implemented in a wide variety of settings. Familiarity with the methodology underlying the framework and code implementation is a great plus, as it helps to adapt the approach to evolving conditions during outbreaks. The translational use of nowcasts in different settings is enhanced by close collaboration with decision-makers. Because non-technical people may not be familiar with the model’s intricacy or the interpretation of its outputs, bridging this gap through accessible language, clear visualizations, and ongoing dialogue is essential to ensure that the nowcasts are informative and actionable for public health professionals.

Our current model has some limitations. We do not explicitly account for seasonal dynamics, alternative data streams (e.g., Google Trends, social media activity), or human mobility, all of which can be extended in the future and may offer additional predictive power. The current approach aggregates cases by epidemiologic week, which may be sensitive to intra week variability in reporting. Implementing daily nowcasts could mitigate this issue by capturing day-of-week effects.

In conclusion, our findings highlight the feasibility and utility of real-time dengue nowcasting for outbreak monitoring and response. Stable reporting systems and strategic model design, such as pooling parameters for small populations, are key to optimizing nowcast accuracy. These insights provide a foundation for expanding nowcasting applications to other infectious diseases and settings, and for enabling earlier, evidence-based public health interventions.

## Data Availability

Simulated data and code to run the nowcasts and perform model validation are available on GitHub (https://github.com/CDCgov/PR_nowcast_2024).

## Appendix

### Nowcasting by Bayesian Smoothing (NobBS) models

Using a modified version of the NobBS approach, we estimated the total number of cases eventually reported for epidemiological week (EW) *t*, denoted as *N_t_*. This quantity represents the sum of observed dengue cases reported with a delay of *d* weeks, defined as the difference between the report week and the onset week, such that as 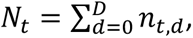, where *D* is the nowcasting window specific to each nowcasting week. To determine *D*, we first calculated the distribution of reporting delays (in weeks) from onset date to report date for cases reported in the most recent week, and set *D* as the 95^th^ percentile of these delays, following prior evidence that this is an optimal cutoff for nowcasting (Lopez et al. 2024). This cutoff ensures the window captured nearly all reporting delays while excluding extreme outliers, and its re-calculation each week allowed for dynamic changes in reporting during the epidemic.

We assume that *n_t_*_,*d*_ follows a negative binomial (NB) distribution

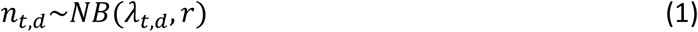

Here, we used an alternative parameterization of the NB distribution, where the mean is the expected number of cases, *λ_t_*_,*d*_, and the dispersion parameter, *r,* represents how overdispersed the case count is relative to a Poisson distribution (Stoklosa, Blakey, and Hui 2022). The expected number of cases, *λ_t_*_,*d*_, is then modeled on the log scale as

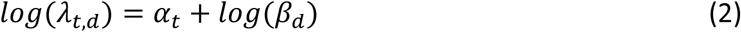

Here, *α_t_* represents the true epidemiological signal at time *t*, while *β_d_* represents the probability of reporting with a delay of *d* weeks.

To capture the temporal evolution of the epidemiological signal, we model *α_t_* as a random walk

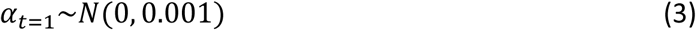

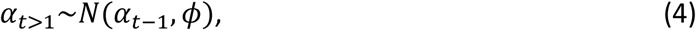

where *ϕ* is the variance of the random walk process. This framework enables estimation of the number of cases *N_t_* by integrating uncertainty in both reporting delays and the underlying epidemic trend.

We used Bayesian inference methods for parameter estimation, specifically Markov Chain Monte Carlo (MCMC) sampling to obtain posterior distributions. Weak priors are assigned to the variance parameter *ϕ* ∼ *Gamma*(0.01, 0.01) and dispersion parameter *r* ∼ *Gamma*(0.01, 0.01). The prior for the delay probabilities *β_d_* are a Dirichlet-distributed vector

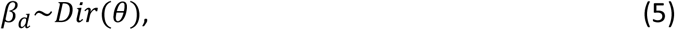

where *θ* = [0.1, 0.1, 0.1, …, 0.1]*^D^*^+1^ is a vector of concentration parameters. The Dirichlet distribution is a natural choice because it defines a probability vector where all elements are nonnegative and sum to one, while *θ* controls how concentrated or dispersed the reporting delay probabilities are around their mean. We set *θ* as a vector with all elements equal to 0.1, reflecting the expectation that reporting delays are predominantly concentrated in a single week, rather than being uniformly distributed across the nowcasting window.

The model is coded in Stan (Stan Development Team 2022). To estimate the posterior distributions, each model was run with 4 chains, 2000 iterations and a 50% burn-in period. Model convergence is assessed by the Gelman-Rubin statistic *R̂* (Gelman and Rubin 1992). All analyses were conducted in R 4.4.0 (R Core Team 2025).

### Baseline model

To establish a simple reference point for evaluating whether a more complex nowcasting method like NobBS offers meaningful improvements, we developed a baseline model where the median estimate is the reported cases in the most recent week preceding the D weeks of nowcasting window, and the variability can be defined through these steps:

1. Define weekly difference distribution: Calculate weekly changes in case counts over the past year as Δ*N_t_* = *N_t_* − *N_t_*_−1_
2. Capture bidirectional variability: Construct a sample distribution using both Δ *N_t_* and −Δ *N_t_* to account for increases and decreases in weekly cases.
3. Generate nowcasted cases: For each time step *t+1*, generate nowcasted case counts by adding a random sample drawn from the difference distribution to the most recent observed case count.

We repeated the sampling process to produce nowcasts for weeks *−D* to 0, aligning with the nowcasting window used in the main model.

### Weighted Interval Scores

The Weighted Interval Score (WIS) is a proper scoring rule used to evaluate probabilistic prediction given in terms of quantiles or prediction intervals (Bracher et al. 2021). It balances prediction accuracy with uncertainty representation, rewarding predictions that are both sharp and well-calibrated. WIS is especially useful when the prediction is expressed across multiple intervals, like the 50%, 80%, and 90% prediction intervals.

To compute the WIS, we first compute the interval score (IS), defined as

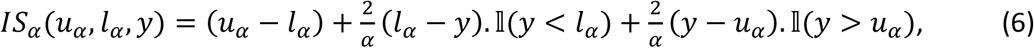

where *u_α_* and *l_α_* is the upper and lower bound of prediction interval *α*, y is the number of reported cases, and l(.) is the indicator function (equal to 1 if the condition is true and 0 otherwise). The interval score penalizes both overprediction, shown as (*l_α_* − *y*) in equation (1), and underprediction, shown as (*y* − *u_α_*) in equation (1), based on how far outside the bounds the observation falls. The weights 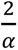 (where *α* is the percentage prediction interval) ensure that narrower intervals (with higher confidence) are emphasized more.

Given the calculated interval scores for a set of *K* prediction intervals, each defined by a confidence level *α_k_*, WIS also considers the absolute error between the observed value and the median forecast, *m*, and finally expressed as

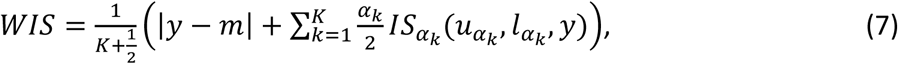

In addition to the absolute error component, |*y* − *m*|, WIS comprises three key components derived from the interval score:

1. Dispersion: The width of each prediction interval, reflecting forecast sharpness. This is computed as the weighted sum of (*u_α_* − *l_α_*) of the interval at level *α*.
2. Underprediction Penalty: Applied when the observed value *y* falls below the lower bound of a prediction interval. It is the weighted sum of (*y* − *u_α_*) across relevant intervals.
3. Overprediction Penalty: Applied when the observed value *y* exceeds the upper bound of a prediction interval. It is the weighted sum of (*l_α_* − *y*) across relevant intervals.

### Sensitivity analysis: understanding 2013 nowcast performance

To understand why the nowcast performance in 2013 differed from other years, we examined the reporting delay distributions for each year. Reporting delays were relatively concentrated around 1–2 weeks in 2009 and 2024, and around 3 weeks in 2010. In contrast, 2013 exhibited highly variable and dynamic reporting delays over time. This variability likely contributed to the increased uncertainty nowcasts observed in 2013. Most high uncertainty estimates occurred during periods of substantial shifts in reporting delays, particularly around EW 10 and from EW 24 to EW 45.

In general, the WIS for 2009 and 2010 was lower for the NobBS model compared to the baseline model, indicating better performance. However, this pattern was not observed in 2013. Notably, when the dispersion component is excluded, the sum of the remaining WIS components for NobBS in 2013 (75.7) remains lower than that of the baseline model (142.0), suggesting that elevated dispersion was the main driver of reduced performance.

Since WIS cannot be directly compared across different datasets, we calculated the relative WIS, defined as the WIS of NobBS divided by that of the baseline model, to evaluate performance over time. In 2009, 2010, and 2024, most relative WIS values by EW were below 1, indicating that NobBS consistently outperformed the baseline model in these years. In contrast, 2013 exhibited the opposite pattern, with many weeks where the relative WIS exceeded 1, meaning the baseline often outperformed NobBS. We also observed that NobBS tended to underperform relative to the baseline during periods of rapid case increases or epidemic peaks. While this behavior was rare in 2009, 2010, and 2024, it was far more pronounced in 2013.

### Appendix Figures

**Appendix Figure 1.**
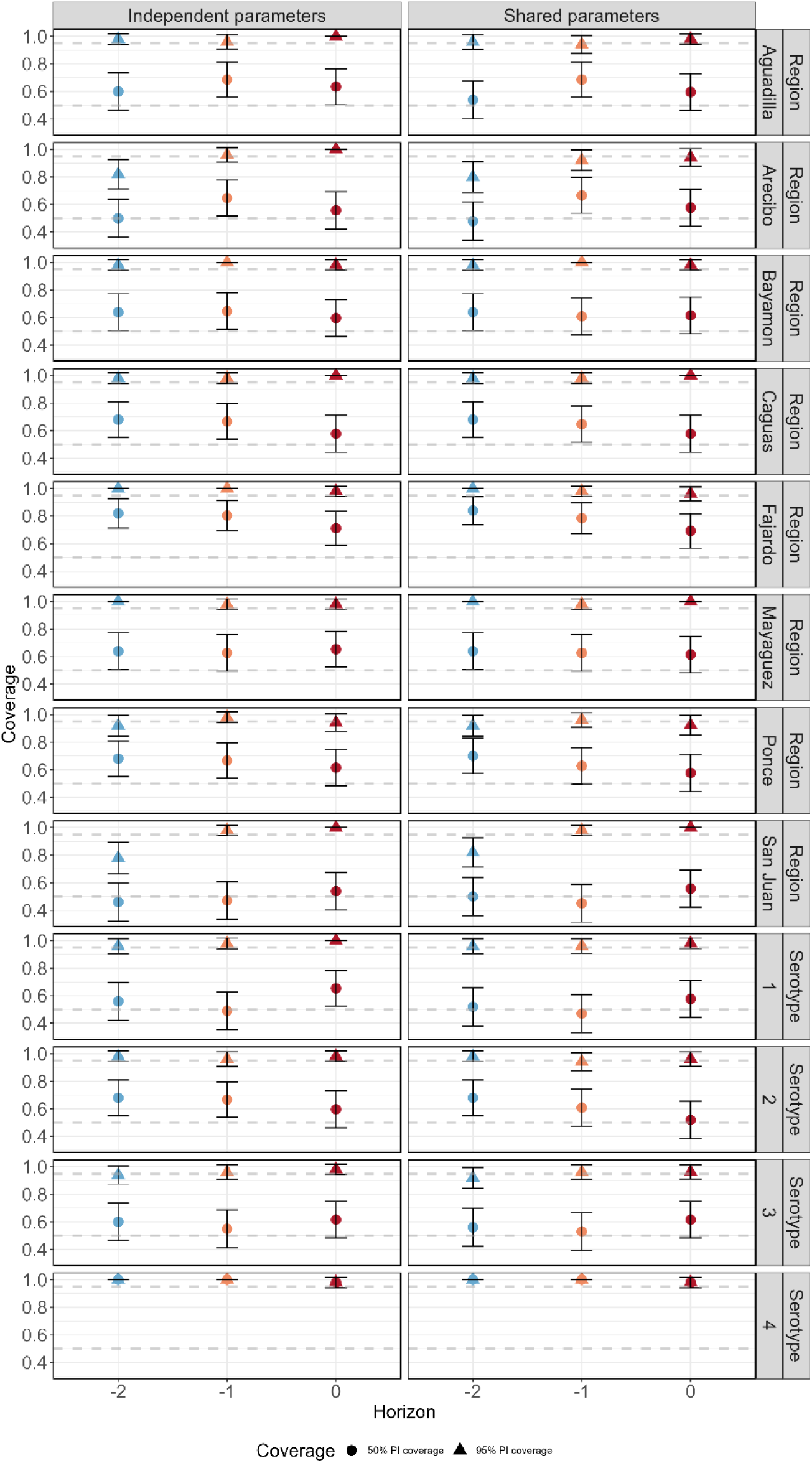
Coverage of prediction intervals (PIs) for NobBS nowcasts across health regions and serotypes during the 2024 dengue epidemic in Puerto Rico. Subpanels show PI coverage for three forecast horizons (−2, −1, 0 weeks) under models with independent parameter (left) and shared parameter (right). Markers indicate 50% PI coverage (circles) and 95% PI coverage (triangles), with error bars representing variability across nowcast weeks. Colors reflect coverage levels, and panels are grouped by health region (e.g., San Juan, Ponce) and serotype (DENV 1-4).

**Appendix Figure 2.**
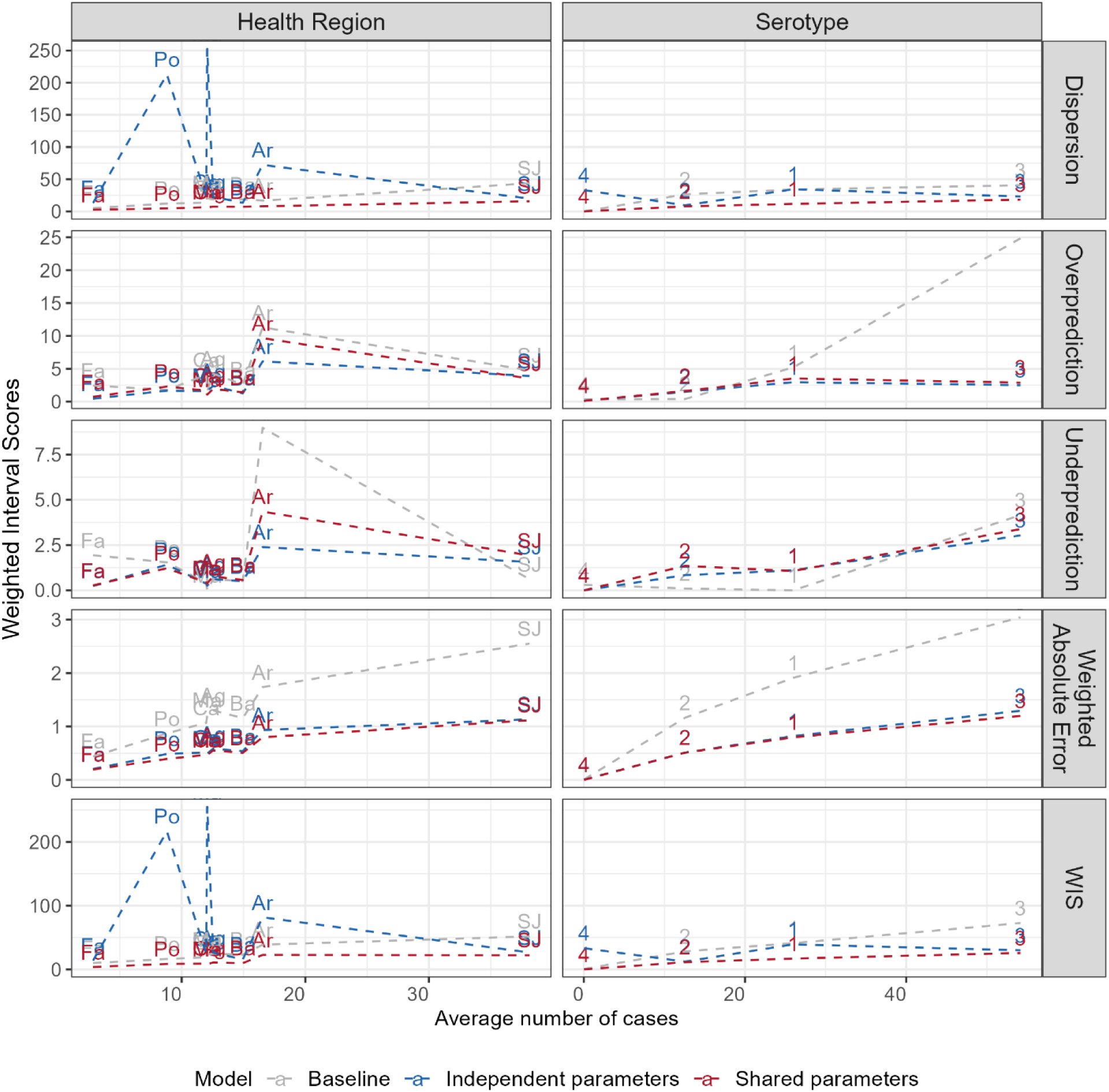
Decomposition of Weighted Interval Score (WIS) for dengue nowcasts in in 2024 in Puerto Rico by health region (left) and serotype (right) over the average number of cases. Rows correspond to different scoring components: Dispersion (top), Overprediction (second), Underprediction (third), Absolute Error (fourth), and total Weighted Interval Score (WIS, bottom), plotted against average number of cases per subgroup. Dashed lines indicate different models: baseline (gray), independent parameter (blue), and shared parameter (red). We would expect WIS to increase along with average number of cases, but this was not observed in the independent parameter model, especially with the dispersion component.

**Appendix Figure 3.**
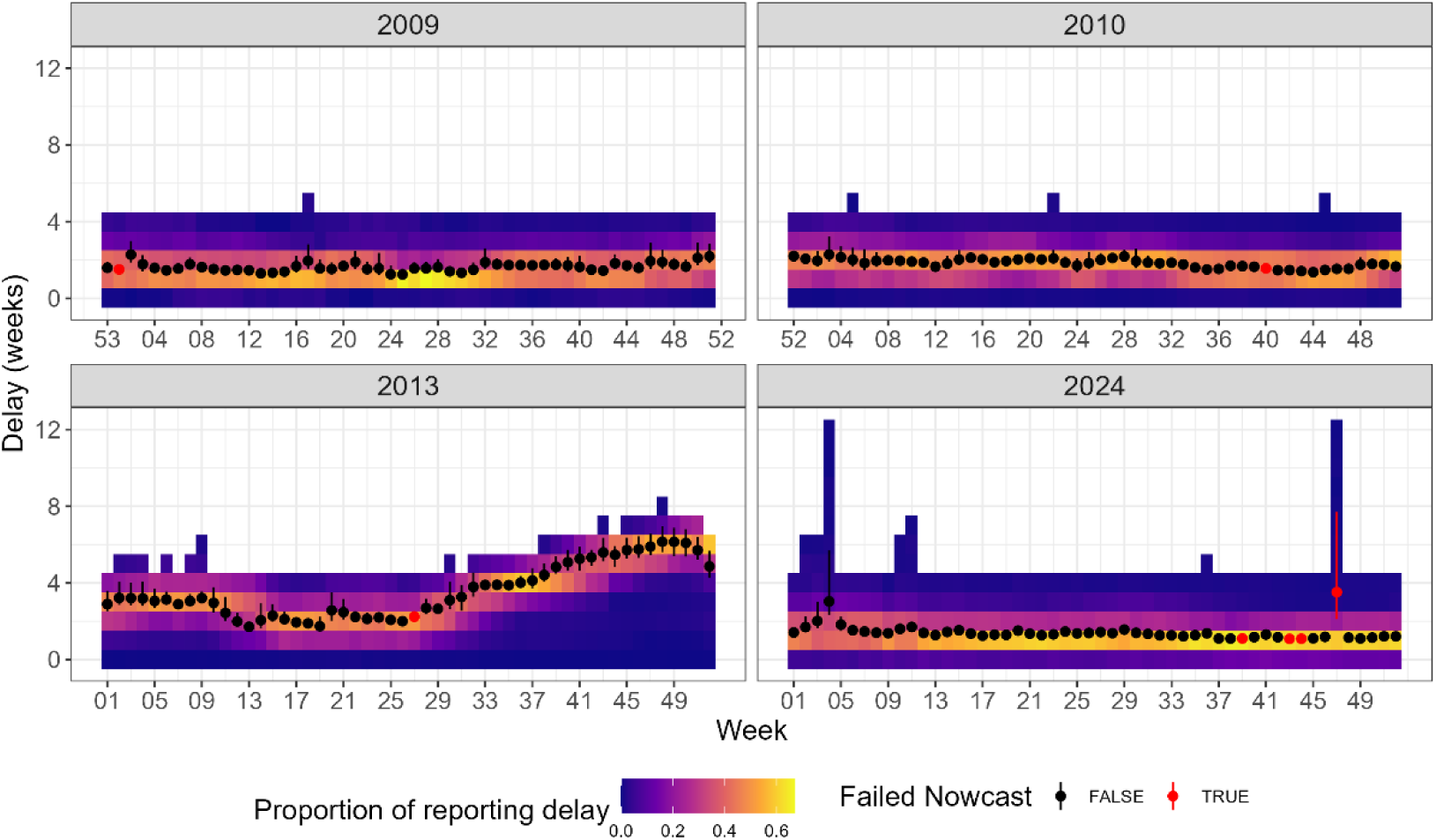
Proportion of reporting delays from island-wide nowcasts for all epidemiological weeks of 2009, 2010, 2013, and 2024. The proportion of reporting delays were calculated from the proportion of reported cases by delay weeks, shown in the legend as a color gradient. The black dots with lines are the median and 95% credible interval estimates of average reporting delays from NobBS, showing alignment with observed delays. Red dots and lines mark the failed nowcast, as determined by log score criteria.

**Appendix Figure 4.**
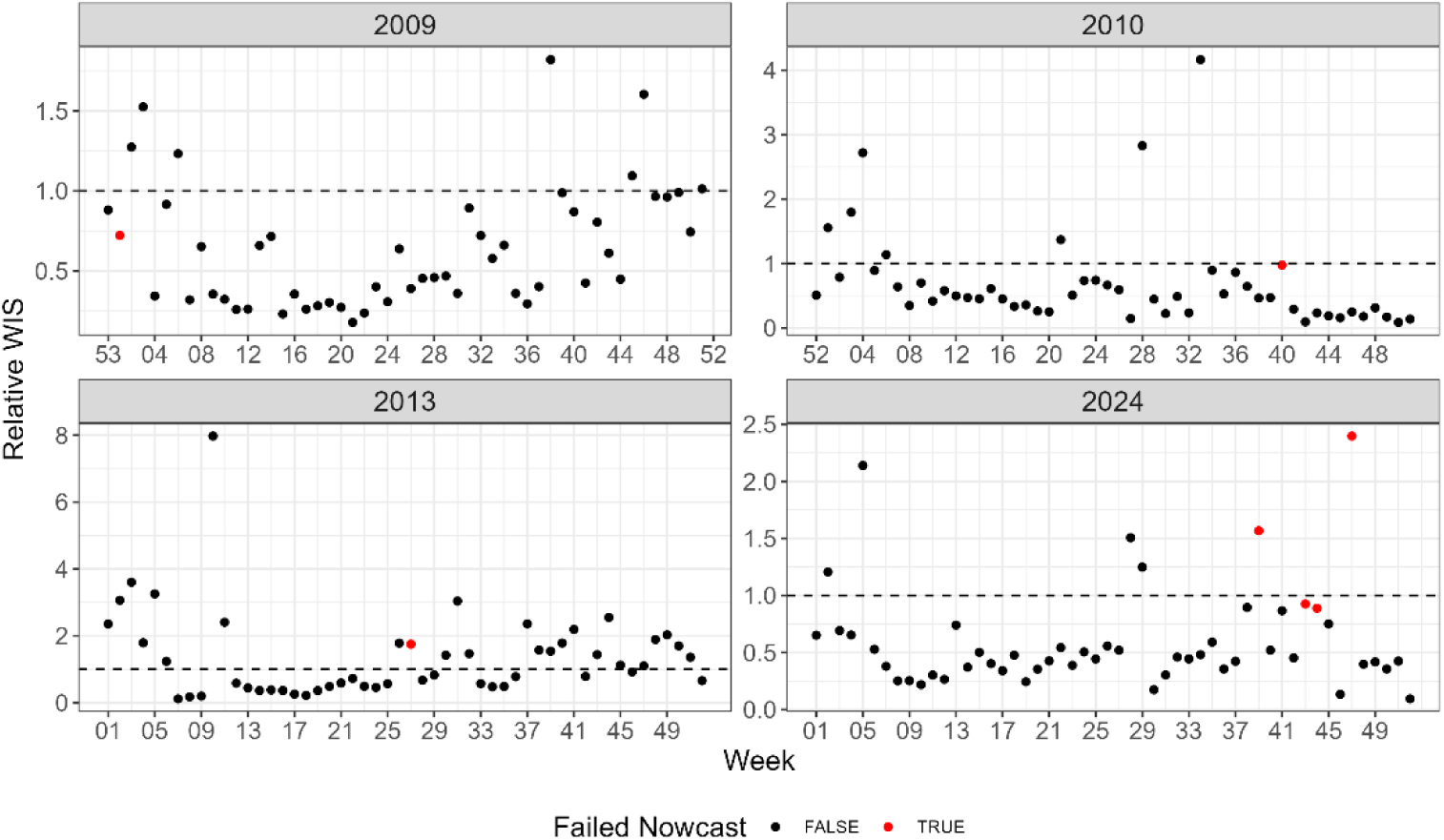
Relative weighted interval score (WIS) for island-wide nowcasts across all weeks in 2009, 2010, 2013, and 2024. Relative WIS was computed as the ratio of NobBS WIS to baseline model WIS, with values below 1 indicating superior performance by NobBS. The horizontal dashed line at one marks the threshold for comparative performance. Red dots denote failed nowcasts as determined by log score criteria.

### Appendix Table

**Appendix Table.** Model performance (WIS with its components and coverage of 50% and 95% prediction intervals) of baseline and NobBS models in year 2009, 2010, 2013, and 2024. Underlined numbers show the lower WIS among the baseline and NobBS. Bold numbers show the coverages that outside of the 50% or 95% target. WIS = weighted interval score, AE = Absolute error, PI = Prediction interval.

| Year | Model | WIS | AE | Dispersion | Underprediction | Overprediction | Coverage 50% PI | Coverage 95% PI |
| --- | --- | --- | --- | --- | --- | --- | --- | --- |
| 2009 | Baseline | 77.51 | 3.72 | 67.46 | 0.51 | 5.82 | **0.8 (0.74 - 0.86)** | **0.99 (0.97 - 1)** |
| 2009 | NobBS | <u>52.88</u> | 1.75 | 41.33 | 1.65 | 8.15 | 0.56 (0.48 - 0.64) | 0.93 (0.89 - 0.97) |
| 2010 | Baseline | 476.39 | 22.08 | 261.00 | 21.24 | 172.06 | **0.74 (0.67 - 0.8)** | **0.88 (0.83 - 0.93)** |
| 2010 | NobBS | <u>255.59</u> | 10.77 | 205.23 | 28.26 | 11.33 | 0.42 (0.34 - 0.5) | 0.96 (0.92 - 0.99) |
| 2013 | Baseline | <u>513.81</u> | 30.44 | 371.94 | 80.99 | 30.44 | **0.69 (0.62 - 0.76)** | 0.94 (0.9 - 0.97) |
| 2013 | NobBS | 608.30 | 17.14 | 532.56 | 18.13 | 40.46 | 0.53 (0.45 - 0.6) | 0.96 (0.93 - 0.99) |
| 2024 | Baseline | 134.13 | 6.21 | 104.35 | 9.28 | 14.29 | **0.63 (0.56 - 0.71)** | 0.95 (0.91 - 0.98) |
| 2024 | NobBS | <u>76.71</u> | 3.28 | 48.59 | 6.19 | 18.64 | **0.37 (0.3 - 0.45)** | **0.88 (0.83 - 0.93)** |

## Disclaimer

The opinions expressed by authors contributing to this journal do not necessarily reflect the opinions of the Centers for Disease Control and Prevention or the institutions with which the authors are affiliated.

